# Clinical Characteristic of a Haitian Stroke Cohort and a scoping review of the literature of stroke among the Haitian population

**DOI:** 10.1101/2022.12.15.22283300

**Authors:** Axler Jean Paul, Jude Hassan Charles, Gandhi Marius Edwitch Gedner, Richardson Roche, Wislet Andre, Garly Rushler Saint Croix, Gillian Gordon Perue

## Abstract

**Background:** There is staggering evidence of stroke care disparities in Low- and Middle-Income countries compared to developed countries. Haiti like those countries suffer from lack of resources for acute stroke management. To our knowledge, we conducted the first study reporting the epidemiological profile of the Haitian population presenting with stroke symptoms in the largest academic hospital of the nation.

**Methods:** This is an observational study conducted over a period of five months from April to August 2021 in the Internal Medicine Department of the State University Hospital of Haiti including 51 consecutive patients suspected to have acute stroke. Descriptive statistical analysis was conducted. A scoping review of the literature on Haitian population stroke was also conducted.

**Result:** More than 50% of our patients are in the age range [19-65] years, 96.1% are older than 35 years. Mean age at presentation was 61 years, predominantly female (64.74 %). Severe motor deficit was more prevalent 96 %, with presenting NIHSS of 12 on average. Only 15% of patients (8/51) had a CT scan during their hospitalization. The majority were delays with a median time to CT of 84 hours after symptoms onset. About 80% of those with complications took more than 24 hours to arrive at the hospital after the onset of symptoms. There was a significant association between the modified Rankin Score and the occurrence of complications (F=6.33; p=0.016). 11% of the patient had complications with a mortality rate of 3.9%. NIHSS score has a very highly significant association with the Glasgow score (p<0.001) where an inverse proportional correlation was measured (r=-0.7; p<0.001) and a proportional correlation with the Rankin prediction score (r=0.3, p=0.04).

**Conclusion:** Stroke affect the most economically active portion of the Haitian population and there is a salient lack of equipped healthcare facilities and appropriate treatment for stroke management in Haiti. Urgent assistance in both personnel and infrastructural resources dedicated to stroke care is needed. Neurological assessment based on NIHSS and Rankin score should be systematic in stroke Evaluation.

## Introduction

Haiti has the highest incidence of stroke in the Caribbean and Latin American region with 176 per 100,000. This incidence is two times higher than the neighboring country the Dominican Republic, ^1^ and represents the first cause of admission to the emergency room. ^2,3^ However little is known about the epidemiology of Stroke in Haiti. In the Global Stroke Statistics 2019 report^4^, Haiti was noted to have a low stroke mortality rate, but the authors acknowledge that the data is more than 18 years old. In a report from world health organization in 2021^5^, 31% of mortality in Haiti can be accounted for by cardiovascular events. Acute stroke treatment is time dependent^6^ including intravenous thrombolysis, mechanical thrombectomy or anticoagulation reversal depending on the stroke subtype. Early management has been shown to promote better outcomes. In Haiti, there are no data available on the time of presentation of stroke patients to the hospital, and the use of the national institute of health stroke scale (NIHSS) score to measure stroke severity is not systematic. Our study aims to describe the clinical characteristics of stroke patients in the largest hospital in Haiti, to highlight challenges associated with stroke management in Haiti including delays in presentation, imaging acquisition and treatment. We include a scoping review of the literature about stroke among Haitian populations.

## Methodology

This is an observational cohort pilot study and a scoping review of Stroke in Haiti, conducted over a period of five months from April 2021 to August 2021 in the Internal Medicine Emergency Department of the State University Hospital of Haiti (HUEH), the main teaching hospital of the Faculty of Medicine of the Haitian State University.

All stroke cases received and admitted to the Internal Medicine emergency department were included in the study. Inclusion criteria were: Stroke patients with complete neurological assessment with Glasgow, NIHSS and Rankin score. Patients referred to us did not often have these parameters, as neurological evaluation with Rankin score and NIHSS is not systematic in other internal medicine departments in the country. During this period, we identified 51 consecutive cases with a clinical diagnosis of stroke that were admitted to HEUH. All stroke cases were considered without determining a sample. We consulted the patients’ medical record after their admission to collect the data using the collection form and we follow their evolution including complications during the hospital course until discharges or death.

A data collection form was used to collect data from suspected stroke cases admitted to the emergency department where the anonymity and confidentiality of patients were protected. The data collection form included details on demographic data such as age, sex, address, occupation, level of education and marital status; clinical data such as chief complaint, history and/or associated defects, date and time of symptoms onset, admission date, vital signs, time to computed tomography (CT) scan, length of stay, referral or not; and a last part on neurological deficits including language, motor and sensory deficits, NIHSS score on admission, modified Ranking score, Glasgow score and state of consciousness as well as outcomes such as whether there were complications, death or recovery. The NIHSS score was used to categorize stroke into mild (1-4), moderate (5-15), severe (16-20), and grave (>20). The Rankin score was used to categorize stroke outcome as favorable (0 to 1 point) or unfavorable (2 to 5 points) depending on the newly considered mRS dichotomization ^7^. Arrival time considered as the time of arrival to hospital after stroke onset was classified into 5 modalities: <3 hours; [3-6]; ]6-12]; ]12-24]and >24 hours after symptom onset.

The data were then entered into Excel and exported to SPSS 26 for processing and analysis. A descriptive analysis of demographic and clinical parameters was performed and presented in table and graph form. Correlation analyses were performed between the NIHSS score and the variables Age, Time of Arrival, SBP, DBP, Glasgow Score, Rankin Score, and length of stay in hospital. Multiple regression analysis was used to specify the forms of significant correlations according to the threshold set. ANOVA test was used to verify associations, Student t test for comparison of means, and Levene test for homogeneity of variances. The significance level was estimated for p<0.05.

A scoping review of the literature was conducted (Table 2). The authors developed a search in PubMed/Medline, EMBASE, Scopus, clinical trials.gov and the Cochrane Library along with several grey literature sources. The searches were conducted from circa 1997 to January 2022. Search was limited to English and French languages and human subjects only. Both subject headings and text searches for terms “ischemic stroke”, “cerebrovascular disease/stroke”, “hemorrhagic stroke”, “brain ischemia”, “acute stroke”, “acute ischemic stroke”, “intracranial occlusion”, or “arterial occlusion”, “subarachnoid hemorrhage” and “Haiti”, “low to middle income countries”, and “Haitian populations”.

## Results

### Demographics

Our study focused on 51 patients admitted for stroke to the internal medicine department, given no dedicated neurology service, at the Haiti State University Hospital from April 2021 to August 2021. Nearly 2/3 (64.74%) of our patients were women, and the mean age was 61.60±16.13 years. More than 50% of our patients are in the age range [19-65] years, 96.1% are older than 35 years, and there is a predominance of women in all age groups. However, males have a slightly higher mean age (62.94 vs. 60.88 years) (Figure 1).

**Figure 1:**
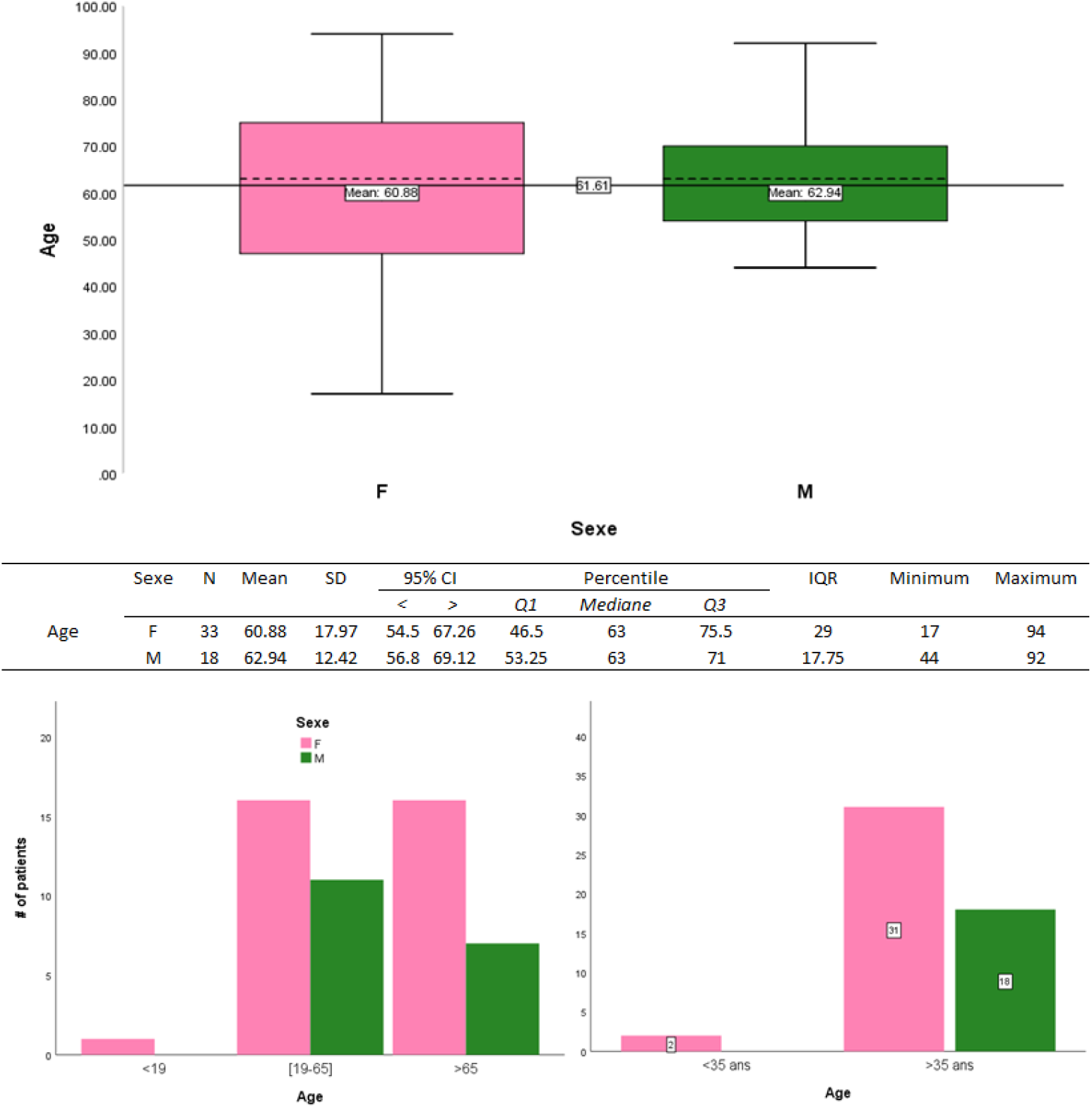
Bivariate analysis between age and sex for our 51 patients.

The district of Port-au-Prince and Carrefour (the closest to the HUEH) were the most represented, with 47% and 25% of patients respectively. Stroke patients coming to the hospital are mostly retailers (49%)

### Clinical Presentations

The 3 main chief complaints were: Alteration of consciousness 18 (35.29%), hemiplegia 10 (19.60) % and aphasia 13 (25.5%). Findings of neurological examination are described in Table 1.

**Table 1:** Clinical characteristics of patients presents with acute stroke symptoms to HEUH

|  |  |
| --- | --- |
| Demographics | Findings |
| Age (Mean +/- SD) | 61.60±16.13 |
| Sex (%Female) | 64.74% |
| Chief Complaints |  |
| Aphasia | 25.50% |
| AMS | 35.29% |
| Hemiparesis | 19.6 |
| Co morbid medical illnesses |  |
| Hypertension | 65% |
| Diabetes | 4% |
| Atrial fibrillation | Unknown |
| Heart Disease | 2% |
| Epilepsy | 2% |
| None | 27% |
| Average arrival Time | 75.14 hours |
| NIHSS (average) | 12 points |
| Time to CT (average) | 84 hours |
| GCS | 11.47 |
| Average Length of Stay | 10 days |
| Outcome |  |
| mRS 0-2 | 74.50% |
| mRS 3-5 | 21.60% |
| mRS 6 | 3.90% |

Aphasia was present in 25.5% of the cases; dysarthria in 29.4%, facial paralysis in 66.7%, a motor deficit in 96.1% of the cases with right sided deficit in 41.1% and left sided 49.1%; and a sensory deficit in 23.5% of the cases. Note that about 10% come on referral from outside small urban clinics. Nearly 2/3 of the patients (65%) were known to have hypertension and 4% were diabetic. The mean arrival time to hospital after symptom onset was 75.14 hours. The NIHSS score for 36/51 patients was on average 12.58 points (Table 1). We found that 7.8% of patients with mild stroke, 41.2% with moderate stroke, 13.7% with severe stroke. 74.5% of the patients had a modified Rankin Score at baseline between 0 to 2 (Table 1).

### Outcomes

Only 15% of patients (8/51) had a CT scan during their hospitalization. The majority were delays with a median time to CT of 84 hours after symptoms onset. The most common finding was that of ischemic findings in one of the middle cerebral artery territories. Patients spent about 10 days in hospital after admission, 6 cases of complications from aspiration pneumonia and 2 deaths were recorded (3.9% in hospital mortality rate). The patients with complications were mostly older than 65 years (83.3%) with a mean age of 69.33±25.48 vs 60.5±14.58 for the uncomplicated cases, the difference was not significant (t=1.25; p=0.21). We observed that 80% of those with complications took more than 24 hours to arrive at the hospital after the onset of symptoms, on average 129.33±105.09 vs. 66.11±106.63 for the uncomplicated cases, the difference is also not significant (t=1.34; p=0.18). All cases of complications were in the category of moderate stroke according to the NIHSS score, the mean score was 17.33±14.46 vs 12.15±5.85 (t=1.28; p=0.20) and the variance is in homogeneous according to Levene’s test F=9.80; p=0.004. However, no association was found (F=1.65; p=0.20). Two thirds (66.7%) of patients had a moderate Glasgow score. (Table 1) There was a significant association between the modified Rankin Score and the occurrence of a complication (F=6.33; p=0.016). The modified Rankin score was higher in patients with complications with a score of 3 or more vs. 0 to 2 in the patients without complications, and the difference was significant (t=2.51; p=0.016). It should be noted that the variance in this group is not homogeneous when considering the Levene test F (5) =5.98; p=0.019. A modified Rankin score of 0.5 (cut off point) has a sensitivity of 60% and a specificity of 86% to predict the occurrence of a complication with an accuracy of 76% (AUC=0.76). (Fig: 2)

**Figure 2:**
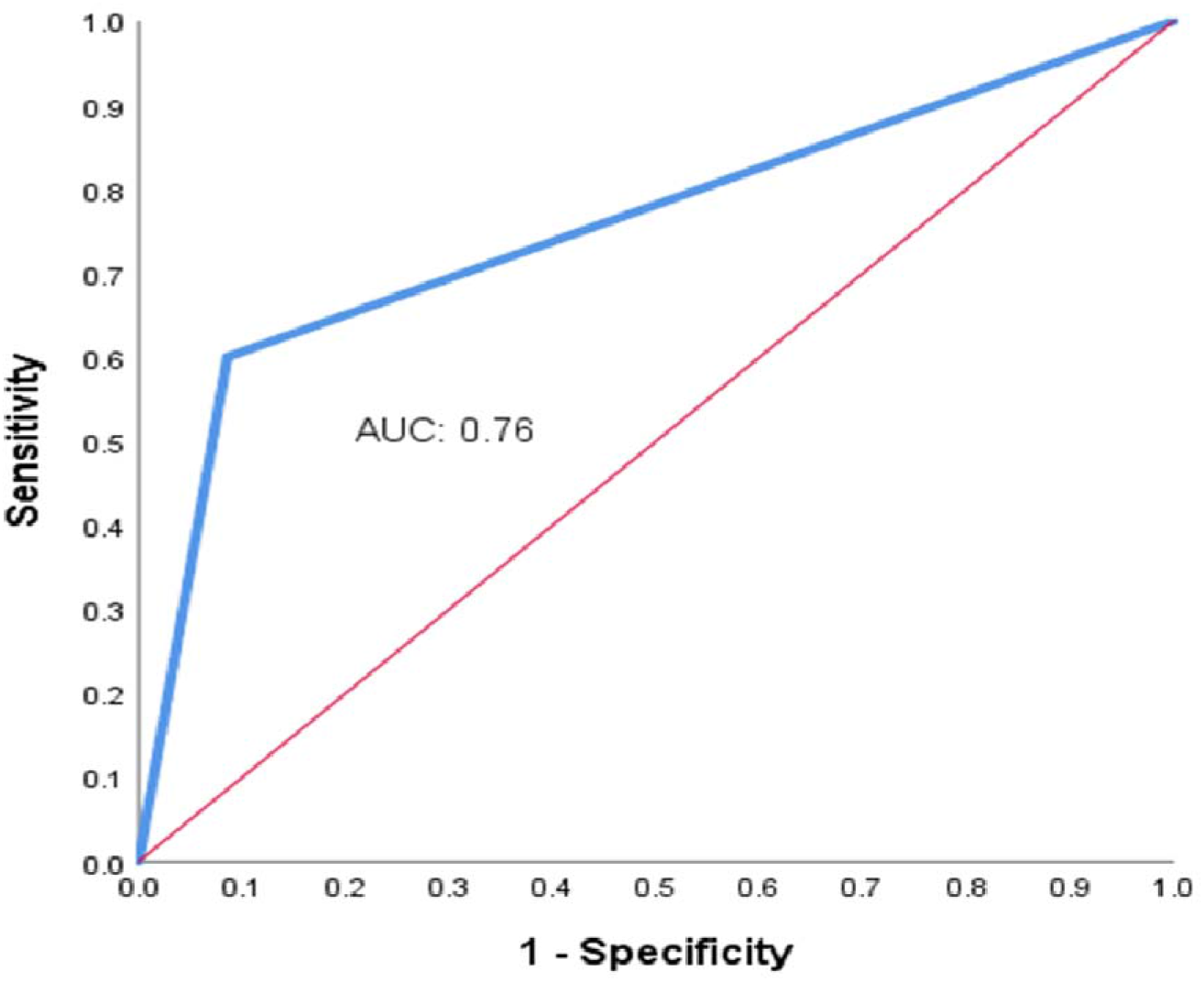
ROC curve analysis to assess the sensitivity and specificity of the Rankin score in the risk of complications.

## Discussion

Stroke is one of the leading causes of death and physical disability in the world.^8^ In Central America and the Caribbean, Haiti has the highest mortality rate from stroke.^1^ To our knowledge, this is the first study on stroke describing sociodemographic and clinical characteristics of stroke patients in Haiti as well as the scarcity of human and material resources in terms of stroke care. The utilization of stroke-related metric such the use of NIHSS, modified Rankin scores, time to intervention is not systematic even in the largest academic center in the metropolitan area. This leads to a lack of data in regard of stroke care in Haiti mainly due to the absence of trained personals.

For the 51 patients of our study, a predominance of women was observed, with a sex ratio of 1.83/1, and this in all age groups, similar to international studies ^9,10^ where a difference of 55,000 more cases of stroke were found annually in women. ^11^ However, men are slightly older than women (62.94 vs 60.88 years). This is different from international observations where women with stroke are often older than men and the average age of stroke patients is around 70 years. ^11–13^ The mean age of our patients is 61.60 years. Stroke among Haitian patients are occurring on average 10 years earlier when compared to developed countries.^1^ For instance, Abreu et al reported an average age of 70 years,^12^ it was 75.39 years in the study of Soto et al.^14^ The average age of our study is close to some developing countries such as Jordan where Qawasmeh et al found 66.5 years and in Tanzania, Matuja et al found 57.9 years ^13,15^ Almost half of the patients (49%) are retailed merchant operating in the informal market. This demonstrates that Stroke is affecting patients in the economic prime of the working years, which result in a significant economic burden to the society. Moreover, and 65% of our patients are known to be hypertensive, a potentially treatable stroke risk factor. It has been shown that the incidence of stroke is increasing in low-income countries, in contrast to high-income countries. ^16,17^ One explanation for this is the impact of social determinants such as poverty, education, lifestyle, and stress on cardiovascular disease, including hypertension, mainly in blacks. ^14,18,19^ A report of WHO from 2021 revealed that about 1/3 of mortality in Haiti is related to a cardiovascular event.

There is evidence that delay in seeking care compromises management, diagnosis and outcomes in stroke patients. ^20,21^ Factors such as symptom onset, history, gender, and stroke severity according to NIHSS score are known to influence the time to hospital. ^21^ The mean time to hospital after symptom onset in our study was 75.14 hours, or 3.13 days. Approximately 39.2% of patients arrived after 24 hours and more than half or 51% of patients arrived after 12 hours. This is much higher than the data in the literature. Mean time to arrive at the hospital is 21.96 hours in China, and 10.73 hours in the USA. ^17^ While the recommended ideal time is 4.5 hours for the use of intravenous thrombolysis and between 6 to 24 hours for endovascular thrombectomy,^21,22^ treatments that are not available in the country. We observed wide variability in time based on sex (99.16 hours for women vs 7.45 hours for men) and the difference was significant (t=-2.59, p=0.013). Similar findings from the study by Le et al, where women arrived later compared to men. ^21^ Immediate assessment is critical in the management of stroke patients, particularly neurological assessment. Several scores are used to assess the neurological status of stroke patients, including the Scandinavian Stroke Scale (SSS), the Canadian Neurological Score (CNS), the modified Rankin Scale (mRS) at admission, and the NIHSS score. ^23^ The NIHSS score is the most widely used because it allows estimation of the neurological impact, severity, complications, and prognosis of stroke with great simplicity, rapidity, and good sensitivity and reproducibility. ^24,25^ In our study, the NIHSS score, and the Glasgow score were used to assess and categorize stroke severity, the modified Rankin score to establish pre-stroke functional level. Patients who developed complications arrived in 80% of cases after 24 hours from the onset of symptoms (129.33±105.09 hours, p=0.18), all had a moderate stroke according to the NIHSS score (17.33 vs±14.46, p=0.20). Schlegel et al in their work also observed an association between NIHSS score and complications (p<0.001), Garavelli et al made the same finding (p=0.048) and Bovim et al observed that 85.4% of the cases of complications had a delay of more than 24 hours after the onset of symptoms. ^25–27^ The Rankin score was higher in patients with complications (1.20±1.64 vs 0.17±0.70) and the difference was significant t=2.51; p=0.016. The Rankin score was significantly associated with the occurrence of complications in patients with stroke (p=0.016). such as aspiration pneumonia with a sensitivity of 60% and a specificity of 86% (AUC=0.76) (Fig.: 3). Thus, the Rankin score can be used to predict the occurrence of complications in stroke patients as studies have shown that the occurrence of complications after stroke is a poor prognostic factor. ^26^ We observed that the NIHSS score has a very highly significant association with the Glasgow score (p<0.001) where an inverse proportional correlation was measured (r=-0.7; p<0.001) and a proportional correlation with the Rankin prediction score (r=0.3, p=0.04). This means that in the initial clinical assessment of stroke patients the NIHSS score, and the Rankin score can add predictive value regarding what patients will likely develop complications. This has been proven by several previous studies.^23,27,28^

Little is known about the epidemiology of stroke patients in Haiti, which limits the ability to create targeted interventions to improve outcomes. In our scoping review, only 3 studies were identified over a 25-year period (Table 2).

**Table 2:** Scoping Review of the Literature

| Table 2: Scoping Review of the Literature |  |  |  |  |  |
| --- | --- | --- | --- | --- | --- |
| Study | Year | Mean age / % sex | Mean NIHSS Reported | Arrival time | Key Finding |
| Stroke epidemiology among Haitians living in Miami (PMID 16103730) | 2005<sup>29</sup> | 60 Years/ 48% female | 7 | N/A | HTN was the most common stroke risk factor at 87 % |
| Head CT findings at public hospital in rural Haiti | 2017<sup>30</sup> | 60.8 ± 17 years / N/A | N/A | N/A | 36% of stroke were hemorrhagic. AMS and focal deficit most common presentation |
| Protocolized emergency department observation care improves quality of ischemic stroke care in Haiti | 2020<sup>31</sup> | 62 ± 14 years / 59% Female | N/A | 4.9 ± 7.1 Days | Reported mean numbers of comorbidities 0.7 ± 0.6 |

The first was done by Koch et al among the Haitian population living in Miami and so it was not able to provide information on the local resources, stroke treatments and outcomes in Haiti. ^29^ The other two studies were conducted in Haiti. Like our study, the mean age of stroke among these Haitian populations is 10 years younger than for developed countries highlight that stroke affects Haitians at their economic prime.^30,31^ Rouhani also document delays in care including delays to arrival time and CT imaging confirming a prolonged time to treatment. ^31^ Together with our current study, these papers demonstrate the need for ongoing research and stroke interventions to save lives and prevent stroke related disability among the Haitian population.

### Limitations

Our study has a small cohort of only 51 patients, which may limit the generalizability of our study. Unfortunately, due to challenges associated with socioeconomic status and limited stroke systems of care none of patients showed up in follow up appointment which limits long term outcome evaluation. This study will serve as the basis for the realization of other more extensive observations on stroke in Haiti.

## Conclusion

Our study shows that stroke knowledge is limited in both the providers and in the population of Port-au-Prince. There is a salient lack of equipped healthcare facilities and appropriate treatment for stroke management. Factors such as low income, limited education, and long arrival time after onset of symptoms are associated with poor outcomes and are found across the board in our population. Neurological assessment based on NIHSS and Rankin score should be systematic in stroke Evaluation. There is an urgent need for assistance in both human and infrastructural resources dedicated to stroke care in Haiti.

## Data Availability

Dataset used and/or analyzed during the current study is available from the corresponding author on reasonable request.

## Ethic approval

This study has been approved by the local ethic committee LABMES (Laboratory of Ethical Medicine and Societies) of the Faculty of Medicine and Pharmacy of the State University of Haiti, and the academic committee of the Department of Internal Medicine authorized us to have access to the file and to the patients during their hospitalization in order to take the data. Verbal consent was obtained from patients or responsible person of patients.

## Consent for publication

Not applicable

## Available data and materials

Dataset used and/or analyzed during the current study available from the corresponding author on reasonable request.

## Authors Contributions

Axler Jean Paul, Jude Hassan Charles, Gandhi Marius Edwitch Gedner and Gillian Gordon Perue have participated in writing the manuscripts; Axler Jean Paul, Gandhi Marius Edwitch Gedner, Roche Richardson, and Andre Wislet are participated in the data collection. Axler Jean Paul, Jude Hassan Charles, Garly Rushler Saint Croix and Gillian Gordon Perue have participated in the methodology and design of the article. All authors have contributed in the revision and correction of the final version of the manuscript.

